# Quantifying statistical power gains from prognostic modeling in Huntington’s Disease

**DOI:** 10.64898/2026.10.19.26365884

**Authors:** Celia Noguerol, Francesco Brizzi, Antoine Soubret, Peter McColgan, Jeffrey D. Long, Marcelo Boareto

## Abstract

**Background and Objectives:** Huntington’s disease is a rare neurodegenerative disease in which optimizing clinical trial design is essential. Although baseline characteristics are known to predict disease progression, the statistical efficiency gains achievable by formally integrating prognostic predictions into trial analyses have not been systematically quantified. We aimed to benchmark prognostic modeling approaches, quantify the trial efficiency gains from prognostic covariate adjustment, and deliver a ready-to-deploy framework for immediate implementation.

**Methods:** We developed a two-step modeling pipeline using longitudinal Enroll-HD natural history data (NCT01574053) to train nine machine learning algorithms to predict disease progression across five UHDRS endpoints. For this, we used 16 predictive baseline covariates representing stage of disease (baseline score levels), disease specific (CAP, CAG length), demographics and co-medication use that could be quantified in one clinical visit. For Total Function Capacity (TFC), frequently used as a primary endpoint in clinical trials, we also devised a minimal prognostic model based on 6 covariates. Model performance was tested using the placebo arm of the GENERATION-HD1 phase 3 trial (NCT03761849).

**Results:** Models were trained on 16,398 Enroll-HD participants and tested on 260 participants from the GENERATION-HD1 placebo arm. Models that take into account nonlinear relationships between the covariates showed no performance advantage over linear-based models on the test set. For TFC, a minimal six-covariate linear prognostic model performed on par with the full 16-feature model. Adjusting for the prognostic score increased the signal-to-noise ratio across different trial visits by 8–18%, raising statistical power from 80% to up to 90% at a fixed sample size, or, equivalently, reducing required sample size by 15–28% compared to unadjusted analyses (no covariate adjustment). Compared to routinely used covariate adjustments (including baseline score, age, CAG repeat length, and CAP score), adjusting for a prognostic score consistently delivered meaningful additional efficiency gains.

**Discussion:** Prognostic covariate adjustment using a simple six-covariate linear score derived from a single baseline visit yields meaningful gains in statistical power for HD clinical trials, while remaining fully embedded within standard, regulator-accepted analysis frameworks. This approach offers a practical and immediately deployable tool to improve trial design.

## Introduction

Huntington disease (HD) is a fatal, inherited neurodegenerative disorder characterized by progressive motor dysfunction, cognitive decline, and functional impairment. Disease progression can be assessed using clinical scores from the Unified Huntington’s Disease Rating Scale (UHDRS), such as the Total Functional Capacity (TFC), Total Motor Score (TMS), Symbol Digit Modalities Test (SDMT), and Stroop Word Reading (SWR), or the composite UHDRS (cUHDRS) score (Keiburtz et al., 1996, Schobel et al., 2017).

Despite significant advances in understanding HD pathophysiology, no disease-modifying therapies are currently approved. Given that HD is a rare disease and several promising therapeutic candidates are approaching human clinical testing, there is an urgent need to optimize clinical trial designs. This brings multiple challenges, as recruitment is naturally constrained, making large sample sizes difficult to achieve. Furthermore, inter-individual variability in disease progression rates across UHDRS endpoints reduces statistical power, requiring long trial durations or large participant cohorts to detect meaningful treatment effects. Computational modelling and data-driven strategies capable of accounting for participant variability can improve trial efficiency by improving statistical power.

Computational and mathematical modelling have substantially advanced our understanding of disease progression in HD. Early modeling efforts focused primarily on predicting age at motor onset using genetic markers such as CAG repeat length (Andrew et al., 1993; Langbehn et al., 2010; Zhang et al., 2011) and the CAG-age product (CAP) score (Warner et al., 2022). More recently, modeling approaches have expanded beyond motor diagnosis to characterize multi-domain longitudinal decline encompassing functional, motor, cognitive, and biological markers (Ghazaleh, et al., 2021; Ko et al., 2022) and defining disease stages by frameworks like the Huntington’s Disease Integrated Staging System (HD-ISS) (Tabrizi et al., 2022). The availability of large observational platforms, such as Enroll-HD, has facilitated the use of machine learning to identify baseline predictive features and map distinct progression trajectories (Ghazaleh, et al., 2021; Ko et al., 2022; Sun et al., 2019; Mohan et al., 2022).

However, translating predictive progression models trained on observational natural history data to interventional trial settings is complicated by fundamental cohort and protocol differences, and trial effects such as placebo response. Recently, Boareto et al. (2025) systematically quantified these discrepancies by comparing Enroll-HD natural history predictions against placebo arm outcomes from the Phase 3 GENERATION-HD1 trial. While functional decline measured by Total Functional Capacity (TFC) aligned with predicted natural history trajectories, motor and cognitive endpoints exhibited clear placebo responses. Specifically, TMS score showed a placebo effect lasting during the whole active dosing period (up to week 69), whereas cognitive scores (SDMT and SWR) displayed sustained improvements likely driven by practice effects from the frequent trial assessment schedule compared with annual observational visits (Boareto et al., 2025). Consequently, composite measures like cUHDRS are influenced by both placebo and practice effects, highlighting that predictive models based on observational data face generalizability challenges when applied to interventional trial settings, and identifying TFC as a suitable endpoint for translation due to its lack of significant placebo or practice responses.

To make use of natural history data for clinical trial optimization, prognostic covariate adjustment (PROCOVA) offers a statistically rigorous solution. However, the statistical efficiency gains achievable by formally integrating observational prognostic predictions into interventional HD trial analyses have not been systematically quantified. PROCOVA has already been applied to other neurological conditions, such as Alzheimer’s disease and spinocerebellar ataxia type 2 (SCA2). In these applications, incorporating pre-specified prognostic scores into trial analyses achieved meaningful efficiency gains, either improving statistical power or enabling sample size reductions (Devanarayan et al., 2025; Tran et al., 2026). This framework uses observational or historical control data to train a prognostic model that outputs a prognostic score (*P_i_*) representing the i-th participant’s expected outcome under placebo or standard of care (Schuler et al. 2022). Incorporating *P_i_* as a pre-specified adjustment covariate within a standard linear regression (ANCOVA), or mixed models for repeated measurements (MMRM), trial analysis can reduce unexplained outcome variability, thereby improve the signal-to-noise ratio and increase statistical power (Schuler et al. 2022). Crucially, because *P_i_* enters the analysis as one baseline covariate, PROCOVA guarantees strict Type I error rate control, and can greatly conserve degrees of freedom. However, maximizing these efficiency gains in trial settings requires prioritizing clinical endpoints resilient to trial-associated effects - such as TFC - and deriving parsimonious prognostic scores that mitigate overfitting while remaining easily computable from routine baseline assessments.

In this study, we aimed to determine the statistical power gains achieved by prognostic covariate adjustment in HD clinical trials and deliver a parsimonious, deployable prognostic score. Using longitudinal natural history data from Enroll-HD, we benchmarked nine machine learning algorithms across five UHDRS clinical endpoints and evaluated out-of-sample generalizability using the placebo arm of the Phase 3 GENERATION-HD1 trial. Focusing on TFC - typically used as primary endpoint in clinical trials - we derived a parsimonious six-covariate linear prognostic score via an exhaustive combinatorial search. Finally, we quantified the variance reduction, signal-to-noise ratio improvement, and sample size improvements achievable when embedding this simplified prognostic score into standard, regulator-accepted trial analysis frameworks.

## Methods

### Standard Protocol Approvals, Registrations, and Participant Consents

The studies were conducted in accordance with the Declaration of Helsinki. For detailed descriptions of the study protocols, please see previously publications for the Enroll-HD study (ClinicalTrials.gov Identifier: NCT01574053) (Landwehrmeyer et al 2017) and the GENERATION-HD1 study (ClinicalTrials.gov Identifier: NCT03761849) (McColgan et al, 2023).

### Study Cohorts

The natural history data used in this work were generously provided by the participants in the Enroll-HD study and made available by CHDI Foundation, Inc. Enroll-HD is a global clinical research platform designed to facilitate clinical research in Huntington disease. We used data from Enroll-HD (NCT01574053) version PDS 7.

Selection criteria and data handling were done as in the previous publication (Boareto et al, 2025). Selection criteria included individuals with an HD-ISS Stage 2 or 3, and at least one annual follow-up visit up. Follow up visits were restricted to up to 4 years to avoid survivor bias and enables the assumption of linearity in the data over this time window. Only individuals who had all the measurements of the baseline clinical scores and covariates were used in the model (in the Covariate Selection section). This led to less than 4% of potential participants being excluded because of missing covariate measurements. Due to such a low level of missingness, no imputation methods were applied. This resulted in a total of 16,398 individuals being selected.

To evaluate the performance of the prognostic models, we used data from GENERATION-HD1 (NCT03761849) (McColgan et al, 2023). This study enrolled 791 adults, with the following overall inclusion criteria: age between 25 and 65 years, diagnostic confidence level (DCL) = 4, independence scale (IS) score ≥70, CAG length ≥36, CAG-age product (CAP) > 400, body mass index (BMI) between 16 and 32 kg/m2, and weight exceeding 40 kg. For more details, please see study protocol (sections 4.1.1 and 4.1.2) (McColgan et al, 2023). This double-blind Phase III study was designed to assess the efficacy and safety of tominersen and participants were equally allocated to a placebo group and 2 active groups at different dosing regimens. Dosing was stopped after most participants had completed the 69 weeks clinical visit, based on an overall benefit-risk assessment by an independent data monitoring committee. Participants were assessed for two more follow-up visits at week 85 and 101. We selected only from the placebo group (n = 260) for the purpose of the analysis in this manuscript. Dropout rate in the placebo arm of approximately 10% at week 69 when dosing was halted and 23% by the final follow-up visit at week 101 were observed.

Baseline characteristics for the selected Enroll-HD and GENERATION-HD1 cohorts are compared in Table 1. Although baseline differences were present between the cohorts, applying the GENERATION-HD1 selection criteria to the Enroll-HD training cohort (Supplementary Table 1) did not substantially alter the overall predictive results. In fact, this filtering slightly reduced model performance, likely due to the decreased sample size in the restricted training set (Supplementary Figure 1). Consequently, we did not perform additional covariate matching or statistical balancing between the cohorts.

**Table 1.** Baseline characteristics.

| Characteristic | Enroll-HD (N = 16,398) | GEN-HD1 (N = 260) |
| --- | --- | --- |
| *Continuous — Mean (SD); Median (Min-Max)* |  |  |
| Age (years) | 47.6 (13.7); 48.0 (19.0–90.0) | 48.7 (9.9); 49.0 (25.0–66.0) |
| CAG repeat length | 43.2 (3.4); 43.0 (36.0–71.0) | 44.9 (3.5); 44.0 (41.0–58.0) |
| CAP100 score | 93.5 (23.1); 97.1 (20.5–193.2) | 107.2 (10.7); 107.2 (83.8–148.2) |
| BMI | 25.4 (4.8); 24.6 (16.7–43.7) | 23.9 (3.2); 23.5 (15.6–32.1) |
| Baseline TFC | 10.6 (3.1); 12.0 (0.0–13.0) | 10.3 (2.3); 11.0 (2.0–13.0) |
| Baseline TMS | 21.1 (20.1); 17.0 (0.0–120.0) | 28.9 (13.2); 28.0 (4.0–78.0) |
| Baseline SDMT | 33.9 (17.2); 32.0 (0.0–101.0) | 26.1 (10.8); 25.0 (2.0–76.0) |
| Baseline SWR | 72.0 (26.1); 72.0 (0.0–220.0) | 66.3 (18.7); 65.0 (21.0–142.0) |
| Independence Scale | 88.5 (14.5); 95.0 (5.0–100.0) | 86.2 (9.4); 85.0 (70.0–100.0) |
| *Binary — N (%)* |  |  |
| Female | 8,870 (54%) | 113 (43%) |
| Education > ISCED 3 | 8,693 (53%) | 160 (62%) |
| HD-ISS Stage 2 | 2,574 (16%) | 37 (14%) |
| Tetrabenazine use | 1,472 (9%) | 31 (12%) |
| Antidepressant use | 4,205 (26%) | 126 (48%) |
| Antipsychotic use | 3,532 (22%) | 75 (29%) |
| Manifest HD (DCL = 4) | 10,214 (62%) | 260 (100%) |

### Prognostic Modelling

Our approach consists of two steps: First, we fit a hierarchical Bayesian model to the longitudinal natural history data (Enroll-HD) to calculate a specific progression rate for each participant. Second, we trained machine learning models to predict these individual progression rates using only baseline data. The final predictions from these models serve as our prognostic scores.

This two-step approach provides two major advantages. First, the Bayesian model naturally handles missing data and irregular visit schedules. Second, because the machine learning models predict the underlying progression rate (*r_i_*), the resulting prognostic score can project an individual’s expected clinical endpoint (*x_t_*) at any future time point (*t*) assuming the linear relationship *x_t_* = *x*_0_ + *r_i_* ⋅ *t*. While disease progression over the course of the disease (time scale of decades) is non-linear, a linear approximation is reasonable within the typical timeframe of a clinical trial (2-3 years).

### Step 1 - Estimating individual progression rates

To estimate the individual progression rate for each individual *i* across the five UHDRS endpoints, we used a hierarchical Bayesian linear model over the observed clinical scores *x_it_*, where *t* denotes visit time relative to baseline (in years). The model is specified as follows:

Likelihood:

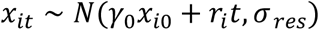

Random effects:

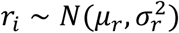

Hyperpriors:

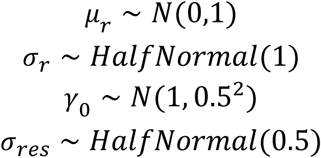

Where the terms and parameters are defined as:

- *γ*_0_: a baseline scaling coefficient that accounts for how strongly the initial score carries forward into future visits. Its prior is centered at 1.
- *r_i_*: the individual (participant-specific) progression rate for individual i, representing their yearly rate of change in clinical score.
- *μ_r_*: the population mean progression rate (fixed effect), representing the average annual change across all participants.
- *σ_r_*: the standard deviation of progression rates across individuals, capturing how many individual trajectories naturally differ from one another.
- *σ_res_*: the residual standard deviation, reflecting the variation in participant scores that is not explained by the participant’s individual progression rate, such as random measurement error, day-to-day fluctuations, or other factors not captured by the linear model.

For each individual *i*, we extract the posterior mean of *r_i_* as the point estimate of their progression rate, along with its posterior standard deviation 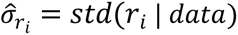, computed across all MCMC samples. In the subsequent ML regression step, each individual is weighted by the inverse posterior variance 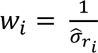, so that individuals whose progression rates are estimated more precisely contribute more to the ML model’s training, reducing the influence of noisy estimates.

### Step 2 - Predicting individual progression rates

Once the individual progression rates were estimated from the Bayesian model, they were used as the target variables for our machine learning models. To account for varying precision across individuals’ trajectories, loss functions were weighted using individuals inverse posterior variance (*w_i_*) previously described.

We benchmarked nine different machine learning algorithms to predict these individual progression rates using only the 16 baseline covariates. Models were categorized based on whether they assumed linear or non-linear functional relationships between the baseline covariates and the target progression rate (*r_i_*):

- **Linear covariate relationship:** Lasso, Ridge, ElasticNet, and Support Vector Machine (SVM) with a linear kernel
- **Non-linear covariate relationship:** Decision Tree, Random Forest, XGBoost, LightGBM, and HistGradientBoosting

For the linear covariate models, features and target values were standardized so that regularization penalties acted uniformly across covariates. Tree-based models used the original, unscaled data since their performance is not affected by feature scale.

### Covariate selection and parsimonious model

The same baseline participant characteristics as described in (Boareto et al, 2025) were evaluated as potential predictive covariates. The following covariates that can be assessed during one clinical visit were included: baseline levels of clinical scores or disease stage (TFC, TMS, SDMT, SWR, IS, HD-ISS, DCL), participant demographics (sex, height, weight, BMI, education level, age), co-medication (such as use of tetrabenazine, antidepressant, or antipsychotic), and disease-specific information (CAG, CAP).

To determine if a simpler, more parsimonious model could perform as well as the full 16 covariates model, we conducted an exhaustive combinatorial search. We evaluated every possible combination of the 16 baseline covariates, resulting in 2^16^ − 1 = 65,535 unique model evaluations. Each combination was fit using a weighted least squares (WLS) linear regression framework, where the loss function was weighted by the inverse variance of the progression rates derived from the hierarchical Bayesian model.

The goal of this search was to isolate a minimal subset of covariates that retained the predictive power of the more complex models while maximizing clinical utility and ease of deployment.

### Model performance

To evaluate model performance using the training dataset, we used a nested cross-validation strategy on the Enroll-HD dataset.

1. **Outer loop**: we used a repeated K-fold cross-validation scheme with 10 repetitions and 10 folds per repetition. This outer loop was used to get an estimate of how well the models generalize
2. **Inner loop:** within each outer fold, a 5-fold cross-validation loop was used to find the best hyperparameters.

After benchmarking the algorithms via nested cross-validation, we trained a final model using the entire Enroll-HD dataset. To select the final hyperparameters for this definitive model, we ran a standard 5-fold cross-validation.

To assess generalizability, the final models were tested on an independent cohort: the placebo arm of the GENERATION-HD1 phase 3 trial. For each individual in the testing cohort, the prognostic score was computed using baseline covariates and the pre-trained model. Predicted change from baseline at each follow-up visit was calculated as the product of the predicted progression rate and the time elapsed since baseline.

The primary validation metric was the coefficient of determination (R^2^) between predicted and observed change from baseline, defined as:

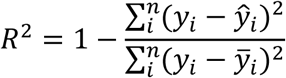

where *y_i_* represents the observed change from baseline for individual i, *ŷ*_*i*_ is the predicted change, and *ȳ*_*i*_ is the mean observed change. This metric directly quantifies the proportion of outcome variance captured by the prognostic score.

When comparing models with differing numbers of covariates, specifically during the exhaustive subset search to identify a minimal prognostic score (see Section “Covariate selection and parsimonious model”), we used the adjusted *R*^2^ to compare models of different dimensionality.

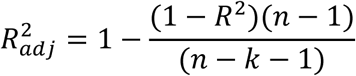

where *n* is the sample size and *k* the number of covariates in the prognostic model.

### Covariate Adjustment

Building on the prognostic score derived in the previous section, we applied the Prognostic Covariate Adjustment (PROCOVA) framework proposed by Schuler et al. (2022). This method involves using the pre-calculated prognostic score (*P_i_*) as an adjustment covariate in the trial analysis. Because the model underlying *P_i_* is trained on external data, and *P_i_* is incorporated as a pre-specified baseline covariate in the trial analysis, this framework helps to control inflation of Type 1 error rate, consistent with regulatory guidance. While the PROCOVA framework is versatile and can be applied to various analysis methods, such as Mixed Model for Repeated Measures (MMRM), we implemented it here using a linear regression model to quantify the reduction in residual variance (σ^2^) achieved by covariate adjustment on the placebo arm. By adjusting for the prognostic score, we account for baseline participant variability, thereby reducing unexplained outcome variance, improving the signal-to-noise ratio, and boosting statistical power. To evaluate the impact of this approach, we compared four scenarios: (1) unadjusted analysis (no covariate adjustment); (2) covariate-adjusted analysis using standard baseline variables (baseline score, age, CAP, and CAG); (3) covariate-adjusted analysis using our model-derived prognostic score; and (4) covariate-adjusted analysis directly using the individual covariates from the prognostic score.

### Sample Size and Power Gain Calculations

Statistical power and required sample size n were estimated using the standard two-sample t-test formula:

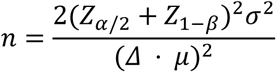

Where:

- *σ*^2^ is the residual (unexplained) variance of the outcome, estimated as *RSS*/(*n* − *p*) where *RSS* is the residual sum of squares and *p* is the number of estimated parameters. This degrees-of-freedom correction ensures that models with more covariates are penalized for the additional parameters consumed, yielding an unbiased variance estimate.

- *μ* is the expected mean change from baseline in the placebo group

- *Δ* is the relative treatment effect, expressed as a proportional reduction in (0, 1].

- (*Δ* ⋅ *μ*) represents the absolute minimum detectable treatment effect

- *α* is the two-sided Type I error rate (significance level, set to 0.05)

- *β* is the Type II error rate (1 − *β* represents the target statistical power, set to 0.80)

- *Z_p_* is the standard normal distribution quantile corresponding to cumulative probability *p* (e.g., *Z_α_*_/2_ and *Z*_1−*β*_).

Rewriting the formula in terms of the signal-to-noise ratio 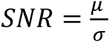 yields:

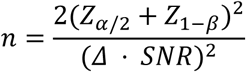

Rather than reporting absolute sample sizes, all quantities are expressed as ratios relative to the unadjusted reference model. Let 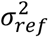 and *SNR_ref_* denote the residual variance and *SNR* under the unadjusted model, and 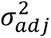 and *SNR_adj_* under the adjusted model. Because all other terms in the formula are held constant, the ratio of required sample sizes simplifies to:

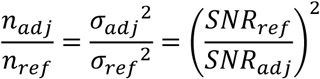

The sample size reduction is then:

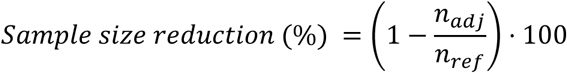

The effective sample size gain (representing the equivalent number of additional participants that unadjusted recruitment would require to match the adjusted analysis) is:

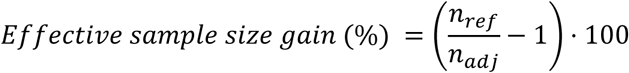

Equivalently, at a fixed sample size *n_ref_*, the statistical power achievable by the adjusted model is:

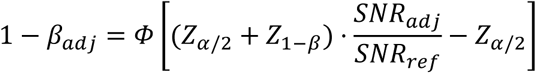

Note that all three metrics (sample size reduction, effective sample size gain, power increase) are directly driven by the ratio 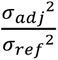. Consequently, introducing any covariate that reduces the residual outcome variance (*σ_adj_*^2^) relative to the reference baseline variance (*σ_ref_*^2^) improves all three outcomes simultaneously. Moreover, comparing these approaches does not require making assumptions regarding the magnitude of a treatment effect (*μ*).

### Software

Figure generation and statistical analysis were performed in Python using NumPy, pandas, and SciPy for data processing, statsmodels for linear regression, and seaborn and matplotlib for visualization. Machine learning models were trained with scikit-learn, XGBoost, and LightGBM, with hyperparameter optimization via Optuna. Hierarchical Bayesian modeling was implemented in PyMC.

### Data Availability

Enroll-HD periodic data set 7 (PDS7) is accessible through the Enroll-HD website (enroll-hd.org/).

For clinical trial studies, qualified researchers may request access to individual participant-level clinical data through a data request platform. At the time of writing, this request platform is Vivli (vivli.org/ourmember/roche/). Up-to-date details on Roche’s Global Policy on the Sharing of Clinical Information and how to request access to related clinical study documents can be found at go.roche.com/data_sharing. Anonymized records for individual participants across more than one data source external to Roche cannot, and should not, be linked because of a potential increase in risk of participant re-identification.

## Results

### Prognostic modeling pipeline

To estimate and predict individual progression rates from baseline characteristics, we applied a two-step pipeline (Figure 1). First, a hierarchical Bayesian model was fitted to longitudinal Enroll-HD data to estimate individual progression rates (rᵢ) for five UHDRS endpoints: TFC, TMS, SDMT, SWR, and cUHDRS. Individual progression rates were estimated using a linear-trajectory model. Although long-term HD progression across decades is non-linear, assuming a linear trajectory is well-justified within our restricted 4-year follow-up window. Second, the estimated progression rates were used as outcomes for supervised machine learning models, trained to predict rᵢ from 16 baseline covariates spanning stage of the disease (TFC, TMS, SDMT, SWR, HDISS, IS, DCL), disease characteristics (CAG, CAP), demographics (Age, Sex, BMI, Education level), and co-medication use (tetrabenazine, antipsychotics, antidepressants) (Figure 1). For more details, please see Methods section and Supplementary Information.

**Figure 1.**
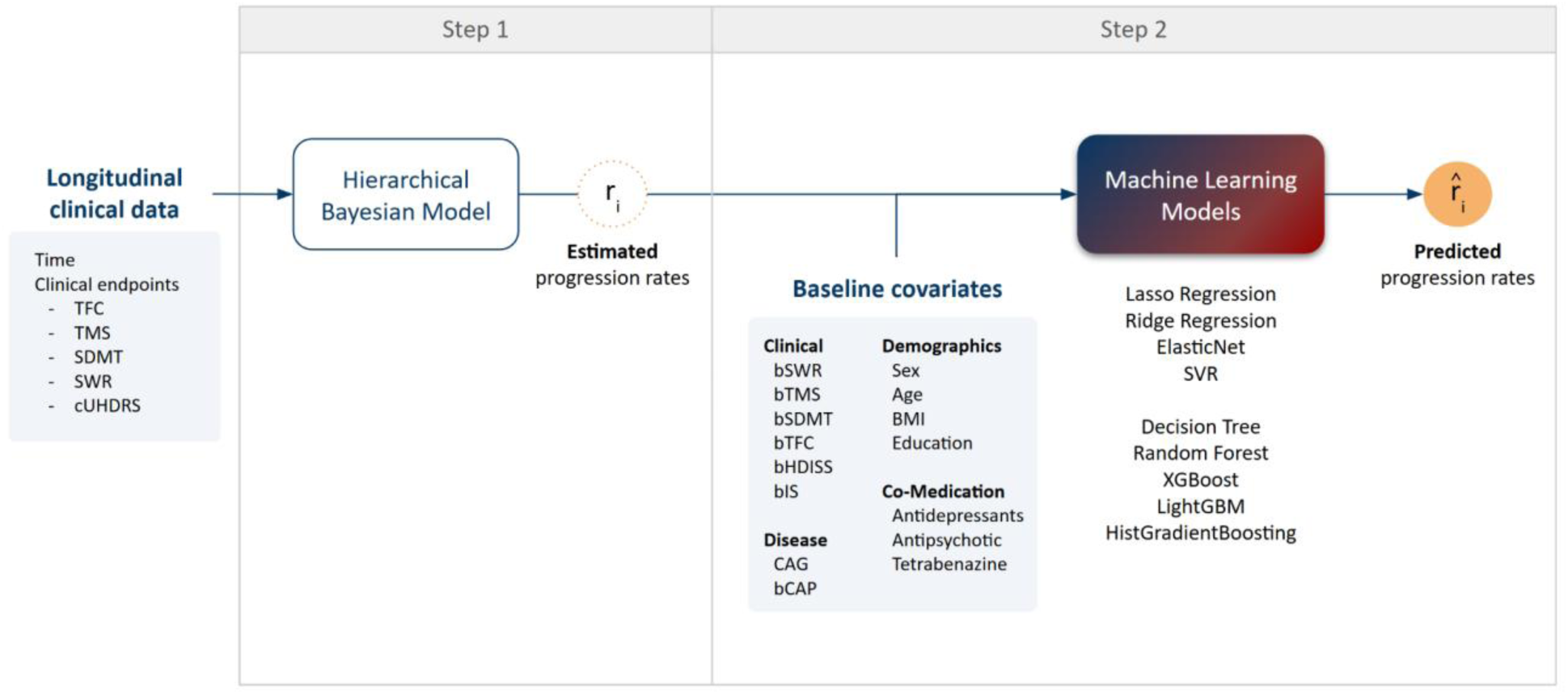
Overview of two-step pipeline. In Step 1, a hierarchical Bayesian linear model is fitted to longitudinal Enroll-HD natural history data to estimate individual-level progression rates (*ri*) across five Unified Huntington’s Disease Rating Scale (UHDRS) endpoints. In Step 2, these estimated progression rates serve as targets to train machine learning algorithms using 16 baseline covariates encompassing disease stage, disease characteristics, demographics, and co-medication. Predicted progression rates are the output of the models.

### Linear and non-linear covariate models perform comparably on testing set

We benchmarked nine machine learning algorithms - four assuming linear relationships (Lasso, Ridge, ElasticNet, SVM) and five capturing non-linear relationships (Decision Tree, Random Forest, XGBoost, LightGBM, HistGradientBoosting) - to predict individual progression rates across all five UHDRS clinical scores (TFC, TMS, SDMT, SWR, cUHDRS). We evaluated model performance by performing a nested cross-validation using Enroll-HD, where 90% of the data was used for training and 10% for estimating performance. While models based on non-linear relationships slightly outperformed linear models during internal cross-validation (Figure 2), this advantage disappeared upon testing in the GENERATION-HD1 placebo arm. On the test set, performance differences between the best linear and non-linear models were minimal, with linear models frequently outperforming non-linear architectures. This shift points to potential overfitting by complex non-linear models on observational training data. Consistent patterns were observed across alternative evaluation time points (Supplementary Figures 2, 3).

**Figure 2.**
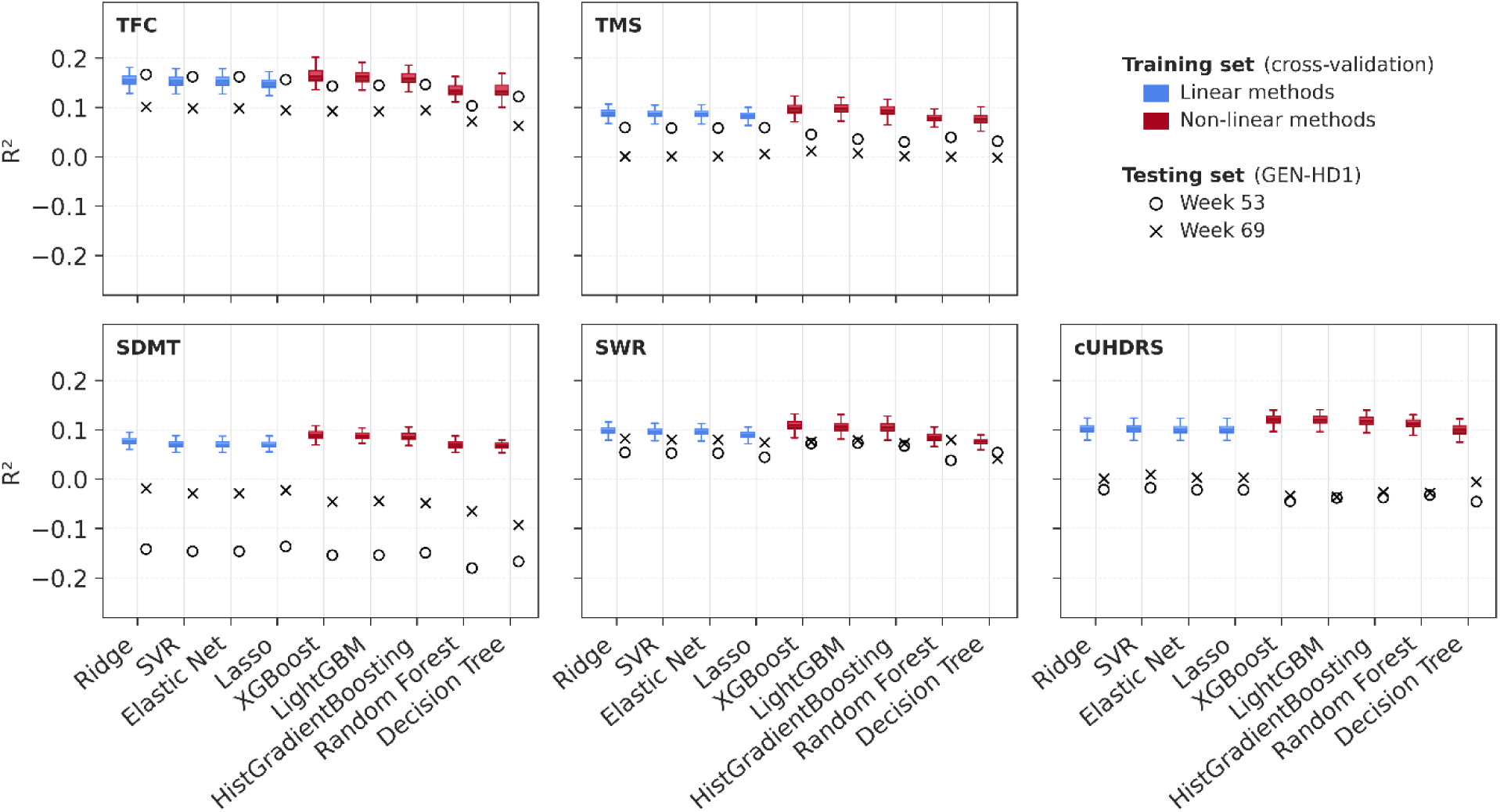
Predictive performance (R^2^) of machine learning models across UHDRS clinical scores. Benchmarking of nine machine learning algorithms - four based on linear relationship between the covariates and outcome (Lasso, Ridge, ElasticNet, and linear SVM) and five based on non-linear relationships (Decision Tree, Random Forest, XGBoost, LightGBM, and HistGradientBoosting) - trained to predict individual disease progression rates (ri) across five UHDRS clinical scores: Total Functional Capacity (TFC), Total Motor Score (TMS), Symbol Digit Modalities Test (SDMT), Stroop Word Reading (SWR), and composite UHDRS (cUHDRS). Models were trained on longitudinal natural history data from Enroll-HD using 10-fold nested cross-validation and tested on the GENERATION-HD1 trial placebo arm.

These findings indicate that added model complexity (in allowing non-linear relationship between the covariates and outcome) does not enhance predictive power and highlight the potential confounding influence of trial-specific effects (such as placebo and practice effects) when applying models trained on observational natural history data to interventional settings.

### TFC shows superior predictive generalizability across trial cohorts

Predictive performance varied substantially across the UHDRS endpoints. TFC consistently showed the highest overall performance and concordance between training and testing cohorts. Conversely, predictive performance was lowest for SDMT, particularly in the testing cohort (Figure 2). This reduced generalizability is likely attributable to the pronounced practice effect previously documented in SDMT during the GENERATION-HD1 trial (Boareto et al., 2025)

Given TFC’s predictability and widespread selection as a primary trial endpoint, we prioritized TFC for subsequent analyses to evaluate whether a parsimonious linear-based model could maintain strong predictive capacity.

### A minimal six-covariate prognostic score for TFC

Having established that linear models perform appropriately, we next sought to identify the smallest subset of baseline covariates required to maintain this predictive capacity. Linear machine learning methods (like Lasso) have coefficients subject to shrinkage penalties to avoid overfitting arising from multiple correlated covariates. To isolate an unpenalized, parsimonious model, we conducted an exhaustive combinatorial search across all possible covariate subsets (2^16^ - 1 = 65,535 models) using Weighted Least Squares linear regression. Covariate subsets - ranging from a single baseline predictor to multi-covariate combinations up to the full set of 16 covariates - were systematically evaluated using a linear regression framework. We found that predictive performance plateaued at approximately six covariates, i.e, incorporating additional covariates beyond this threshold yielded negligible gains in predictive performance (Supplementary Figure 4). Furthermore, within this six-feature threshold, a family of distinct covariate combinations showed comparable predictive performance (Figure 3). This equivalence across multiple candidate models is likely to be driven by high multicollinearity and strong correlations among baseline clinical, demographic, and disease-stage covariates. For downstream analyses, we selected the top-performing six-covariate model as our reference parsimonious prognostic score. In this case, the prognostic score for an individual i (*P_i_*), can be calculated using the following equation:

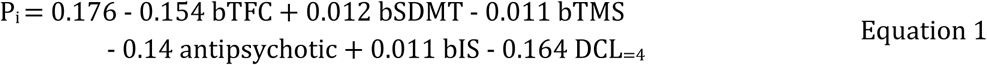

Where:

- *bSDMT*: baseline Symbol Digit Modalities Test (SDMT) score
- *bTMS*: baseline Total Motor Score (TMS)
- *bTFC*: baseline Total Functional Capacity (TFC) score
- *antipsychotic*: a binary indicator variable (coded as 1 if the participant is taking antipsychotic co-medication at baseline; 0 otherwise)
- *bIS*: baseline Independence Scale score
- *DCL*_=4_: a binary indicator variable for diagnostic confidence (coded as 1 if the Diagnostic Confidence Level is 4; 0 otherwise).

**Figure 3.**
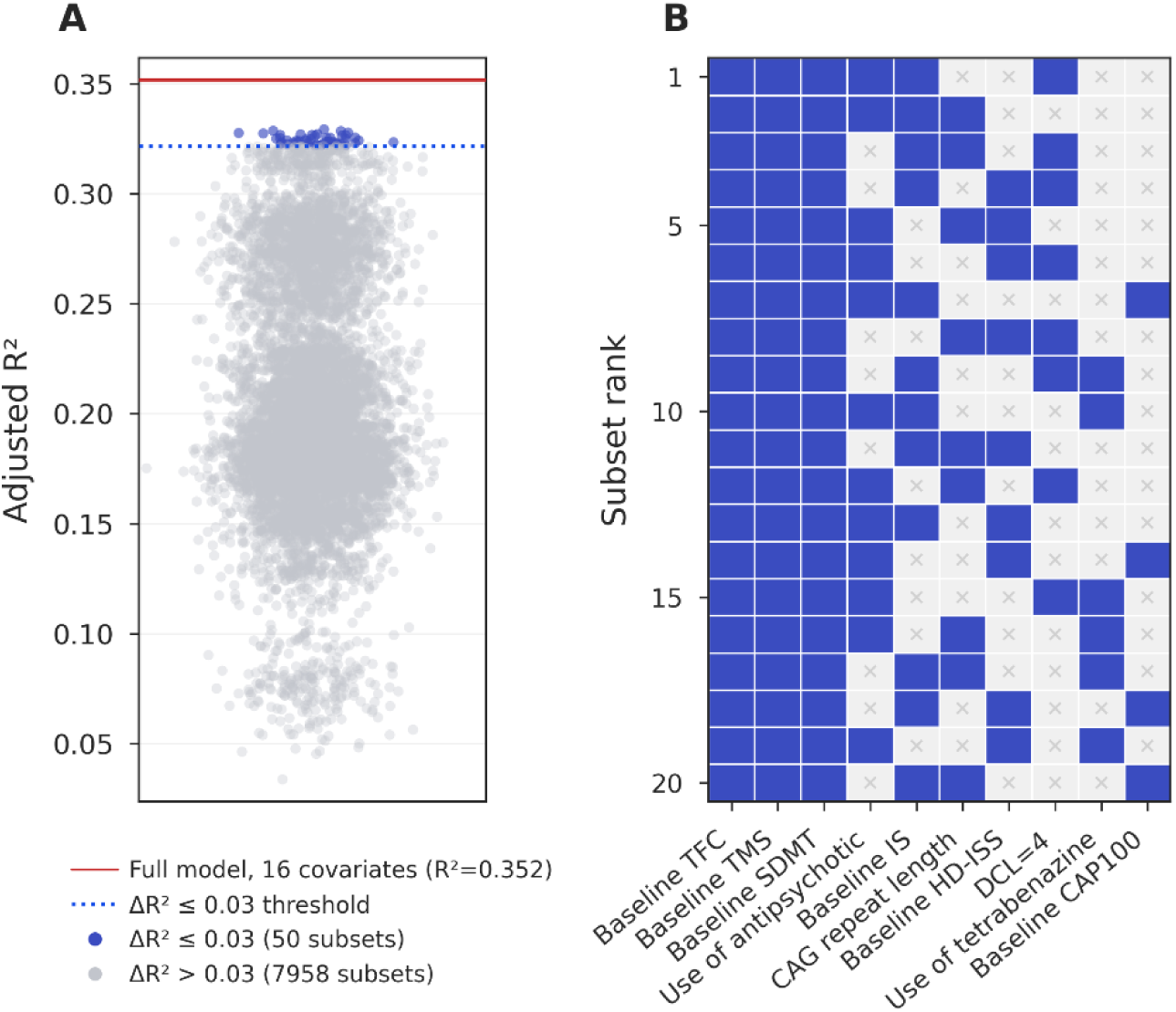
Identification and performance evaluation of a parsimonious prognostic model for TFC. A) R² performance of all 6-feature subsets; the threshold indicates the subsets that have an adjusted R² drop of ≤ 0.03 relative to the full 16-covariate model (totaling 50 subsets). B) Matrix depicts the top 20 subsets out of 50 qualified 6-covariate subsets and their constituent baseline covariates. Covariate coefficient values for each top 20 model are presented in Supplementary Table 2.

### Prognostic covariate adjustment increases statistical power

To evaluate whether incorporating prognostic predictions improves trial efficiency, we implemented the Prognostic Covariate Adjustment (PROCOVA) framework described by Schuler et al. (2022). This method uses a baseline prognostic score (*P_i_*) - derived from historical natural history data - as an adjustment variable in the trial analysis to reduce unexplained outcome noise. To quantify the benefit of this approach, we benchmarked four different analysis strategies on TFC change from baseline using participant data from the GENERATION-HD1 placebo arm:

- Unadjusted analysis (no baseline covariates).
- Standard trial adjustment (adjusting for baseline score, age, CAG repeat length, and CAP score).
- PROCOVA (adjusting for the composite prognostic score *P_i_* derived from Equation 1).
- Individual component adjustment (adjusting directly for the six individual baseline variables that compose *P_i_*).

We evaluated the impact of this framework on predicting changes from baseline in TFC score using participant data from the GENERATION-HD1 placebo arm as a representative HD clinical trial cohort. Linear covariate adjustment incorporating our minimal six-covariate prognostic score achieved a reduction in residual variance for TFC change from baseline ranging across visits from 15% to 27% relative to unadjusted analyses (no covariate adjustment). This variance reduction translated into an 8% to 17% increase in the signal-to-noise ratio across trial visits (Figure 4A). These efficiency gains translated into an increase in statistical power from 80% to up to 90% at a fixed sample size (Figure 4B). Equivalently, this corresponds to an effective sample size gain in the range of 16-40% (Figure 4C), and equivalent sample size reduction in the range of 15-28% (Figure 4D). Importantly, while standard trial covariate adjustment - adjusting by baseline score, age, CAG, CAP - offered consistent gains (SNR increases of 3–10%), our prognostic score consistently provided meaningful performance improvements beyond standard covariate adjustment alone. Similar results were found for all 20 top performing 6-covariate TFC models shown in Figure 3 (Supplementary Figure 5).

**Figure 4.**
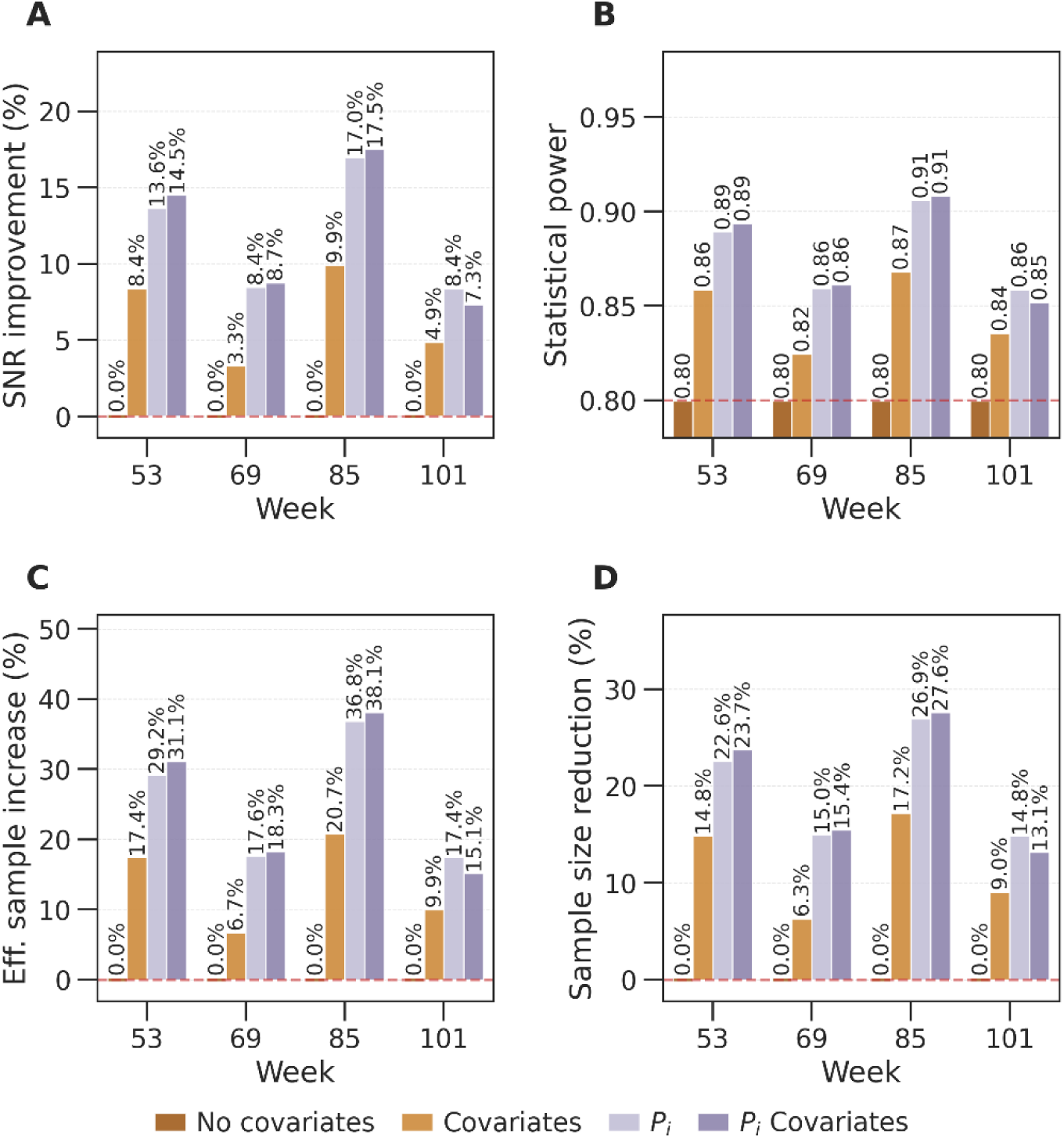
Impact of prognostic covariate adjustment on trial efficiency for TFC. Evaluation of TFC change in the GENERATION-HD1 placebo arm benchmarking four analytical specifications: unadjusted analysis (no covariate adjustment), standard trial adjustment (baseline TFC, age, CAG repeat length, and CAP score), prognostic score alone (P_i_ derived from the reference minimal six-covariate model, Equation 1), and individual covariates from prognostic model (baseline TFC, baseline TMS, baseline SDMT, baseline IS, use of antipsychotic and baseline DCL=4). A) Signal-to-noise ratio (SNR) improvement. B) Statistical power at a fixed trial sample size (alpha = 0.05, two-sided). C) Effective sample size gain, and D) sample size reduction. All analyses were done for the visits at week 53, 69, 85 and 101. Weeks 53 and 69 represent a double-blind period, while weeks 85 and 101 are follow up visits for most participants.

## Discussion

This study evaluates the utility of baseline prognostic modeling to optimize Huntington’s disease clinical trials within standard, regulator-accepted statistical frameworks (May 2023 US Food and Drug Administration (FDA) guidance document). Using observational data from Enroll-HD and the GENERATION-HD1 placebo arm, we show that prognostic covariate adjustment achieves meaningful gains in statistical power, signal-to-noise ratio, and trial efficiency.

We found that while machine learning models that are based on non-linear relationships (e.g., Random Forest, XGBoost) exhibited slightly superior performance on training data (Enroll-HD), this advantage disappeared when evaluated on testing data (GENERATION-HD1). Models based on linear relationships repeatedly matched or exceeded non-linear models in predicting individual progression rates in the testing cohort. This pattern points to potential overfitting of complex algorithms on observational natural history data or to limited generalizability of the models trained on Enroll-HD to trial settings. In this case, parsimonious linear models might be preferable due to their generalizability, transparency, and ease of protocol implementation.

Our results showed differences in predictive performance across UHDRS domains. TFC consistently showed the highest overall performance and good concordance between natural history training and trial testing cohorts. Conversely, cognitive endpoints such as the SDMT showed lower generalizability, characterized by a notable reduction in predictive performance on the testing cohort. This discrepancy can be attributable to score-specific trial effects, particularly the strong practice effects documented in SDMT in GENERATION-HD1, likely due to the more frequent testing schedules during the trial compared to annual testing in Enroll-HD (Boareto et al., 2025). Similarly, motor endpoint (TMS) was shown to be susceptible to transient placebo effects during active dosing regimen. TFC, however, exhibited no evident placebo response in GENERATION-HD1 (Boareto et al., 2025) and given its robust predictability and status as a standard primary endpoint in pivotal trials, it emerged as the most suitable candidate for prognostic adjustment.

Building on this rationale, we derived a minimal prognostic score for TFC through an exhaustive covariate search, finding that predictive performance plateaued using just six baseline variables. Notably, this six-covariate solution was not unique: a broad family of alternative combinations achieved comparable performance, reflecting the substantial multicollinearity among baseline clinical, demographic, and disease-stage measures in HD. This redundancy is practically useful, as it means sponsors are not locked into a single fixed panel of covariates and can substitute variables as needed without sacrificing predictive performance. Integrating our minimal six-covariate prognostic score into trial analysis using linear covariate adjustment translates into tangible operational efficiency. By including the prognostic score (*P_i_*) as an explicit covariate, we observed an increased statistical power from 80% to nearly 90% at a fixed sample size, or equivalent sample size reduction in the range of 15-28% tested across multiple visits and compared to no covariate adjustment. Standard trial adjustment (using baseline TFC, age, CAG and baseline CAP) offered consistent gains (equivalent to sample size reduction in the range of 6-17%). However, our prognostic score consistently provided meaningful performance improvements beyond standard covariate adjustment.

From a practical drug development perspective, this linear six-covariate framework offers analytical flexibility and a compelling, phased strategy for optimizing clinical programs. Because the underlying relationship is linear, investigators can choose either to adjust directly for the six baseline covariates within a standard ANCOVA model or to condense them into a single scalar prognostic score (*P_i_*), yielding comparable reductions in residual variance. Crucially, using *P_i_* consumes only one degree of freedom in the primary analysis model - a meaningful advantage when preserving residual degrees of freedom is essential to maximizing statistical precision. This is often the case for a Phase 2 trial, and it can help convert borderline, inconclusive signals into more confident, interpretable readouts. This same Phase 2 setting also offers a natural opportunity to prospectively test the approach’s predictive performance within a specific trial population before committing to its use in a pivotal study. Once validated in this way, sponsors can pre-specify the prognostic model in Phase 3 protocols with greater confidence, deliberately reducing sample size requirements and thereby shortening recruitment timelines, lowering trial costs, and minimizing overall patient burden.

This framework offers further practical advantages beyond its phased applicability. The identification of a broad family of equivalent six-covariate models provides additional flexibility, allowing trial sponsors to substitute specific baseline variables if certain clinical metrics or co-medication records are unavailable, or part of inclusion criteria, without compromising predictive performance. Importantly, because linear covariate adjustment only uses baseline covariates, these efficiency gains are achieved while preserving Type I error rate control and aligning fully with regulatory recommendations (May 2023 US Food and Drug Administration (FDA) guidance document). Finally, because the minimal score relies on only six variables measurable at a single clinical visit, it can be embedded into standard trial protocols with minimal added logistical burden.

Several limitations of this work warrant consideration. First, our results were tested on a single phase 3 trial placebo arm, which, while representing the largest available placebo dataset in manifest HD, may not fully capture trial-to-trial variations in participant demographics or trial protocols. Second, our study cohorts focused primarily on individuals in HD-ISS Stages 2 and 3; consequently, the generalizability of this minimal prognostic score to early or very advanced disease stages requires dedicated validation. Finally, while our six-covariate score captures part of the predictable progression variability, residual unexplained variability remains, driven by unmeasured environmental, biological modifiers of disease trajectory, or treatment effects.

In conclusion, prognostic covariate adjustment using a simple, six-covariate linear score derived from natural history data offers a practical and powerful strategy to optimize clinical trial design in Huntington disease. By reducing required sample sizes and improving statistical power without inflating false-positive risks, this approach provides an immediately deployable framework to increase trial efficiency, lower patient burden, and accelerate the development of disease-modifying therapies.

## Data Availability

Enroll-HD periodic data set 7 (PDS7) is accessible through the Enroll-HD website (enroll-hd.org/).
For clinical trial studies, qualified researchers may request access to individual participant-level clinical data through a data request platform. At the time of writing, this request platform is Vivli (vivli.org/ourmember/roche/). Up-to-date details on Roche's Global Policy on the Sharing of Clinical Information and how to request access to related clinical study documents can be found at go.roche.com/data_sharing. Anonymized records for individual participants across more than one data source external to Roche cannot, and should not, be linked because of a potential increase in risk of participant re-identification.

## Acknowledgments

Natural history data used in this work were generously provided by the participants in the Enroll-HD study and made available by CHDI Foundation, Inc. Enroll-HD is a clinical research platform and longitudinal observational study for families with Huntington’s disease intended to accelerate progress toward therapeutics; it is sponsored by CHDI Foundation, a nonprofit biomedical research organization exclusively dedicated to collaboratively developing therapeutics for HD. Enroll-HD would not be possible without the vital contribution of the research participants and their families. The authors thank all CHDI/Enroll-HD participants and all the participants in GENERATION-HD1, their families, the site staff, and the trial team, for their time and commitment to the trial.

## Author contributions

Conceptualization: CN, JDL, MB

Methodology: CN, FB, AS, MB

Investigation: CN

Supervision: MB

Writing – original draft: CN, MB

Writing – review & editing: CN, FB, AS, PM, JDL, MB

## Supplementary Information

**Supplementary Table 1.** Baseline characteristics. Comparison between Enroll-HD (when GENERATION-HD1 selection criteria is applied) and GENERATION-HD1 cohorts

| Characteristic | Enroll-HD (N = 5,946) | GEN-HD1 (N = 260) |
| --- | --- | --- |
| *Continuous — Mean (SD); Median (Min-Max)* |  |  |
| Age (years) | 49.5 (9.7); 51.0 (25.0–65.0) | 48.7 (9.9); 49.0 (25.0–66.0) |
| CAG repeat length | 44.5 (3.2); 44.0 (40.0–67.0) | 44.9 (3.5); 44.0 (41.0–58.0) |
| CAP100 score | 106.2 (11.1); 105.7 (77.0–186.3) | 107.2 (10.7); 107.2 (83.8–148.2) |
| BMI | 24.1 (3.4); 23.8 (16.7–32.0) | 23.9 (3.2); 23.5 (15.6–32.1) |
| Baseline TFC | 10.1 (2.4); 10.0 (2.0–13.0) | 10.3 (2.3); 11.0 (2.0–13.0) |
| Baseline TMS | 29.6 (14.3); 28.0 (0.0–89.0) | 28.9 (13.2); 28.0 (4.0–78.0) |
| Baseline SDMT | 25.5 (10.7); 24.0 (0.0–89.0) | 26.1 (10.8); 25.0 (2.0–76.0) |
| Baseline SWR | 61.8 (19.1); 61.0 (0.0–132.0) | 66.3 (18.7); 65.0 (21.0–142.0) |
| Independence Scale | 86.2 (10.4); 85.0 (70.0–100.0) | 86.2 (9.4); 85.0 (70.0–100.0) |
| *Binary — N (%)* |  |  |
| Female | 2,938 (49%) | 113 (43%) |
| Education > ISCED 3 | 2,825 (48%) | 160 (62%) |
| HD-ISS Stage 2 | 941 (16%) | 37 (14%) |
| Tetrabenazine use | 741 (12%) | 31 (12%) |
| Antidepressant use | 1,839 (31%) | 126 (48%) |
| Antipsychotic use | 1,697 (29%) | 75 (29%) |
| Manifest HD (DCL = 4) | 5,946 (100%) | 260 (100%) |

**Supplementary Table 2.**
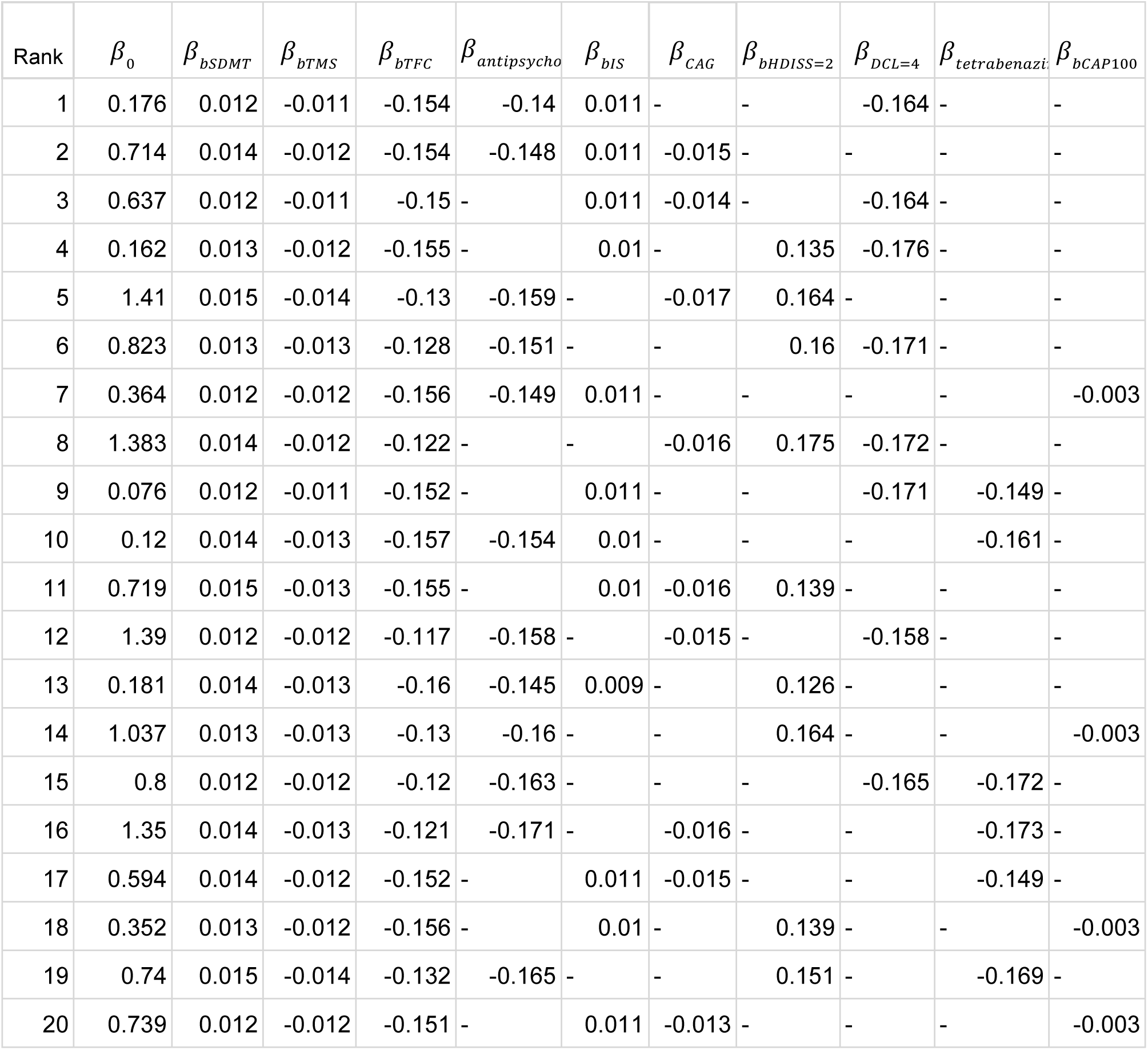
Covariate coefficient of all top 20 top performing 6-covariate models for TFC.

**Supplementary Figure 1.**
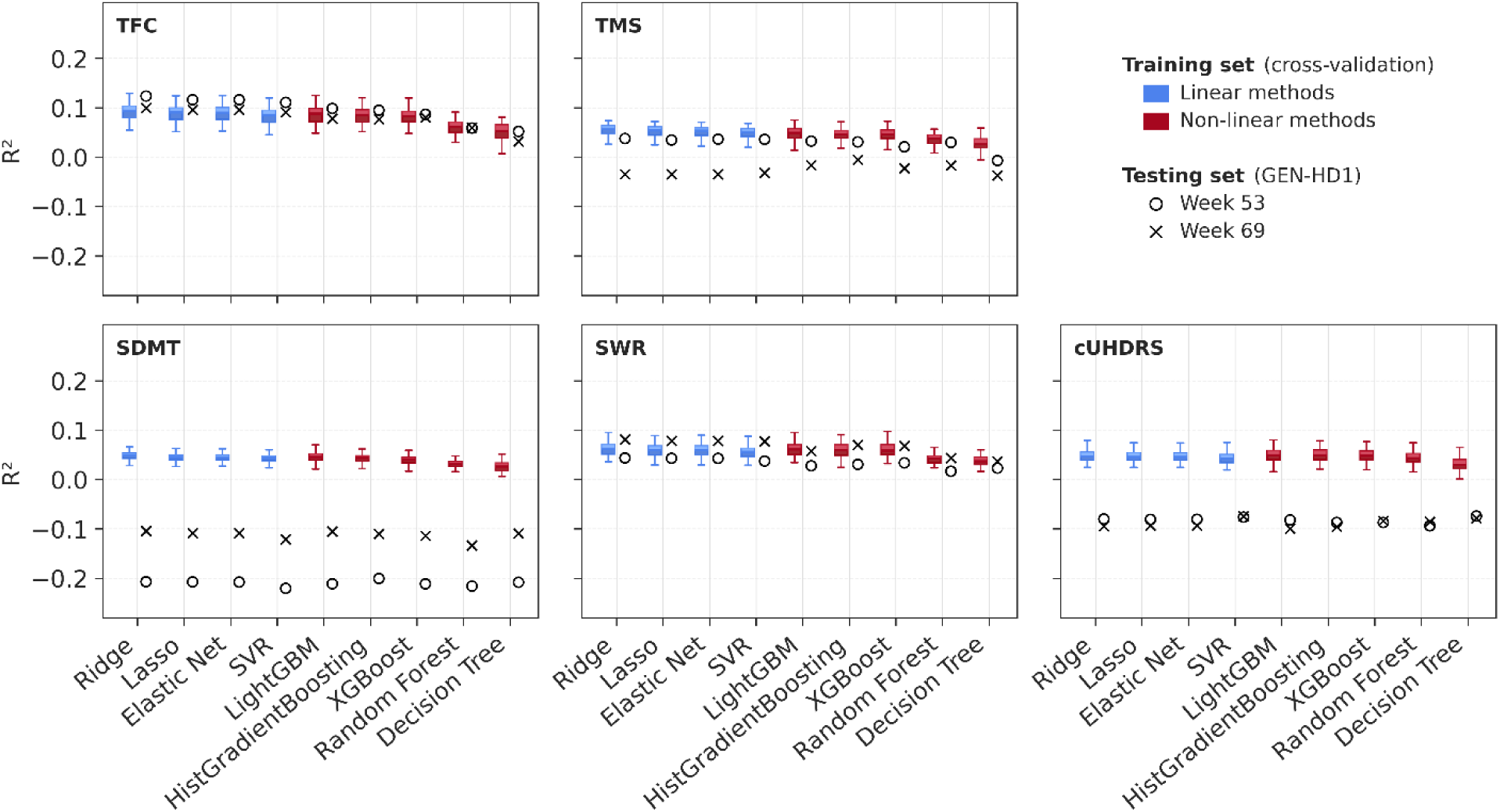
Same as Figure 2 but training done using Enroll-HD with the selection criteria from the GENERATION-HD1 trial.

**Supplementary Figure 2.**
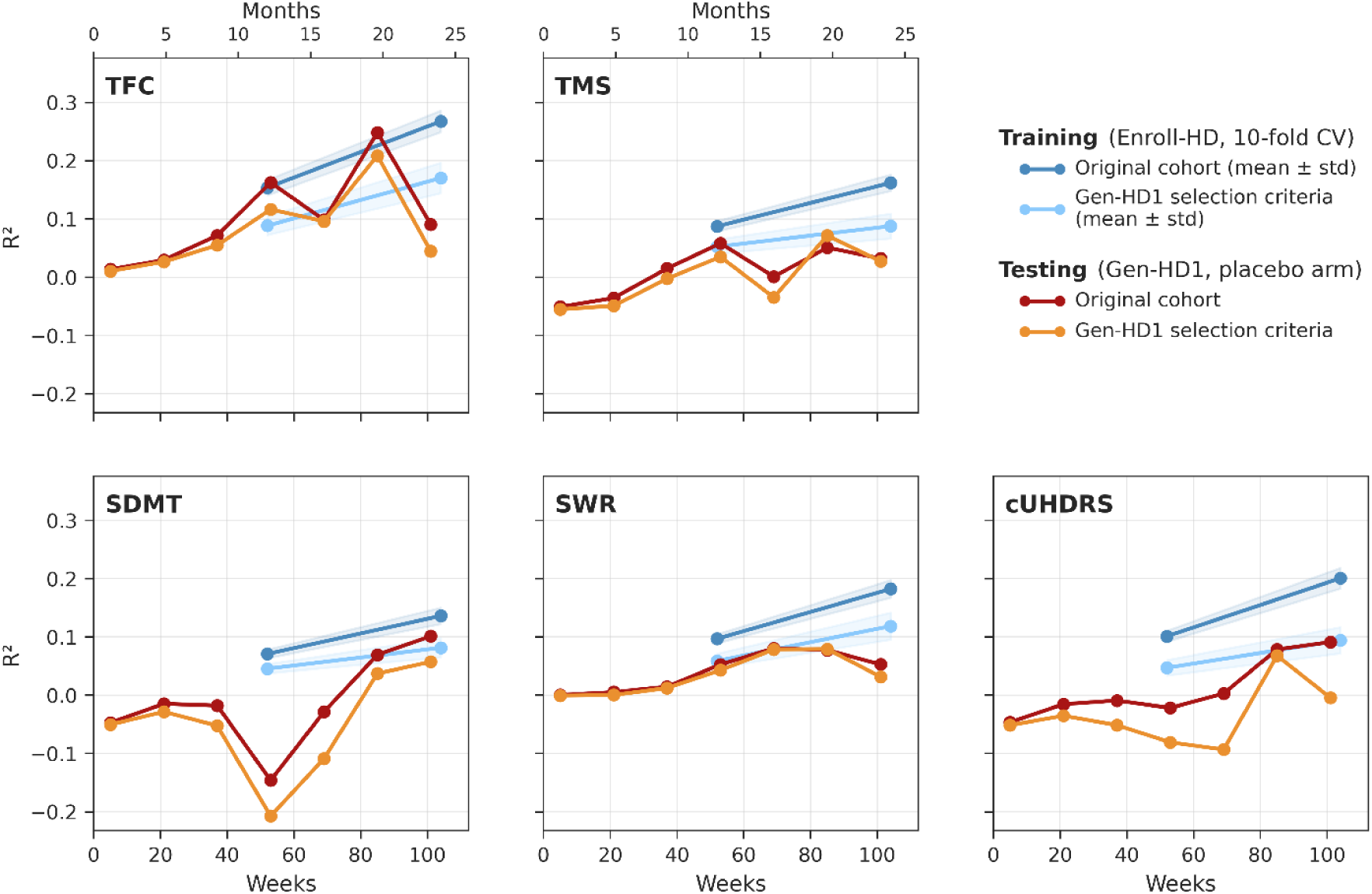
R^2^ performance over time for different clinical endpoints using the Lasso model, comparing training (cross-validation) and testing (GENERATION-HD1 placebo arm) performance. Comparisons are shown using both the full Enroll-HD cohort and the subset filtered by GENERATION-HD1 trial selection criteria.

**Supplementary Figure 3.**
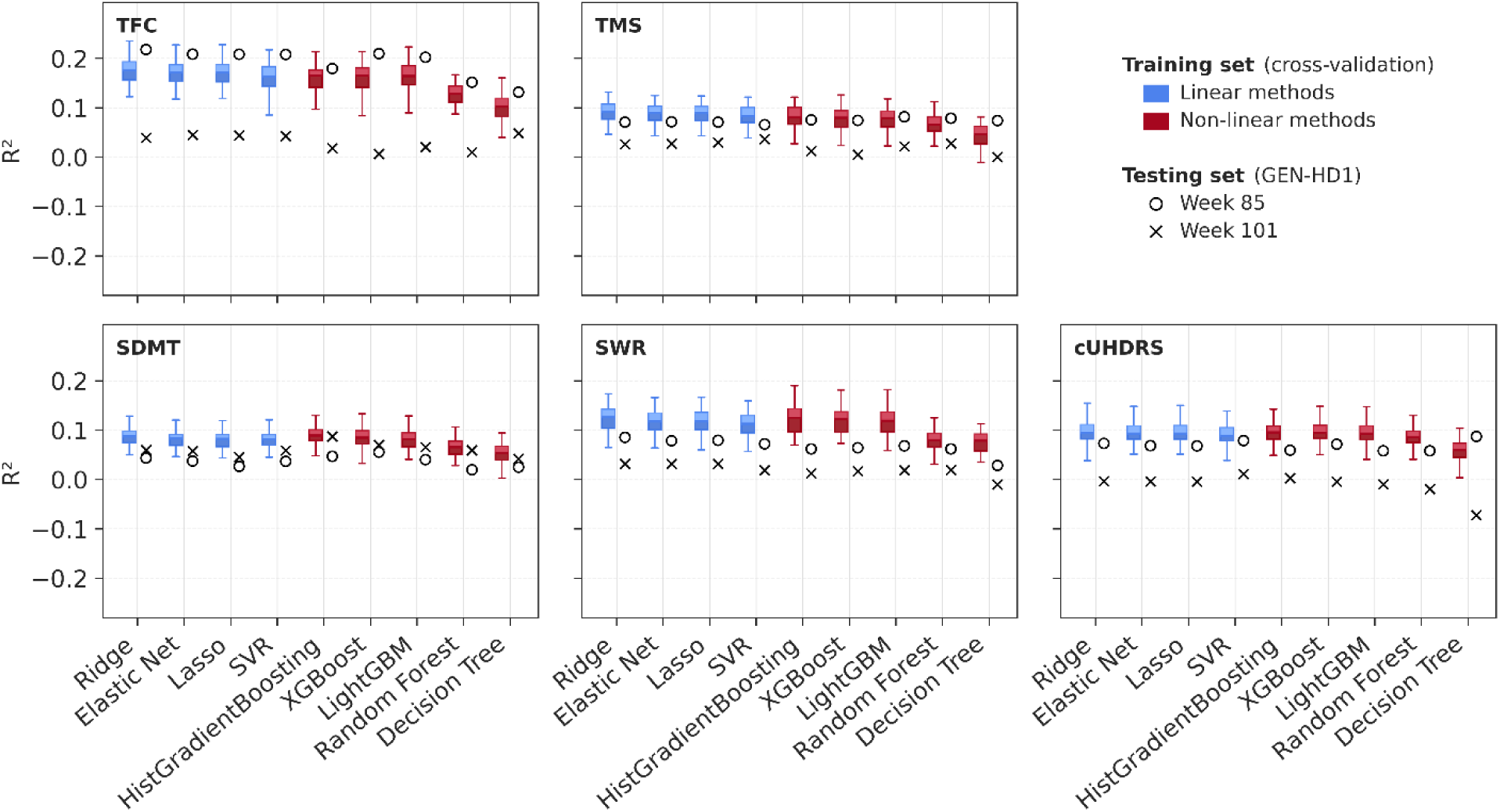
Same as Figure 2 but for training done using Enroll-HD visits at 2 years. Testing was done for GENERATION-HD1 clinical visits at week 85 and 101. It is important to note that for most participants visits at week 85 and 101 were follow-up visits, and not part of the double-blind period of the trial, as the trial was stopped after most participants participated on visit week 69.

**Supplementary Figure 4.**
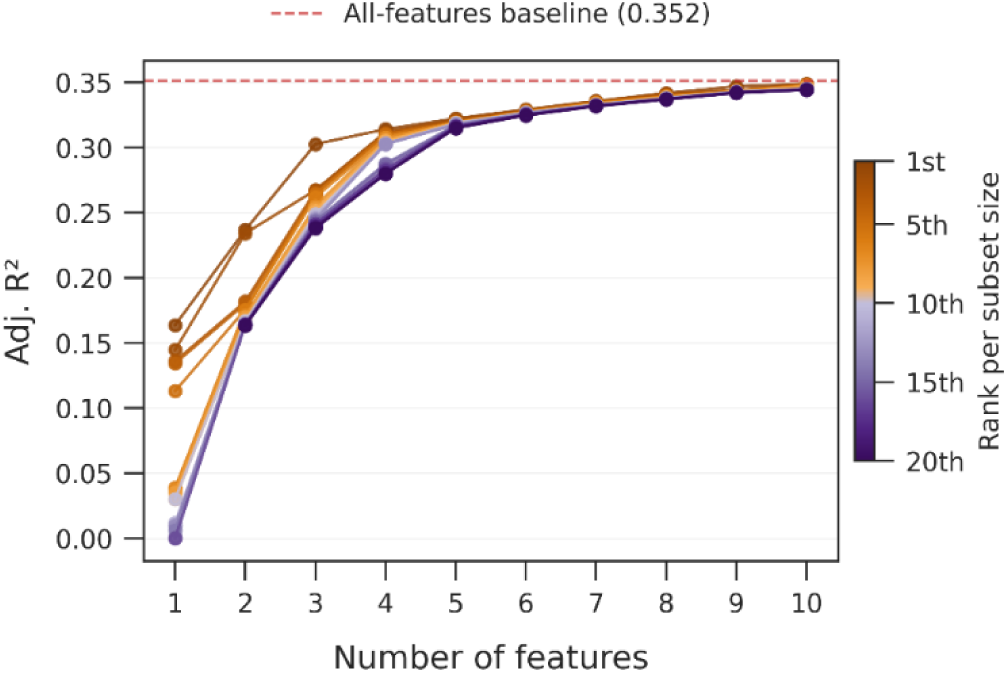
Adjusted R^2^ performance of the top 20 ranked feature subsets (colored by rank) plotted against the number of features selected. Performance varies considerably for smaller features sizes, but converges around 6 features, after which all top 20 subsets achieve comparable R^2^ performance.

**Supplementary Figure 5.**
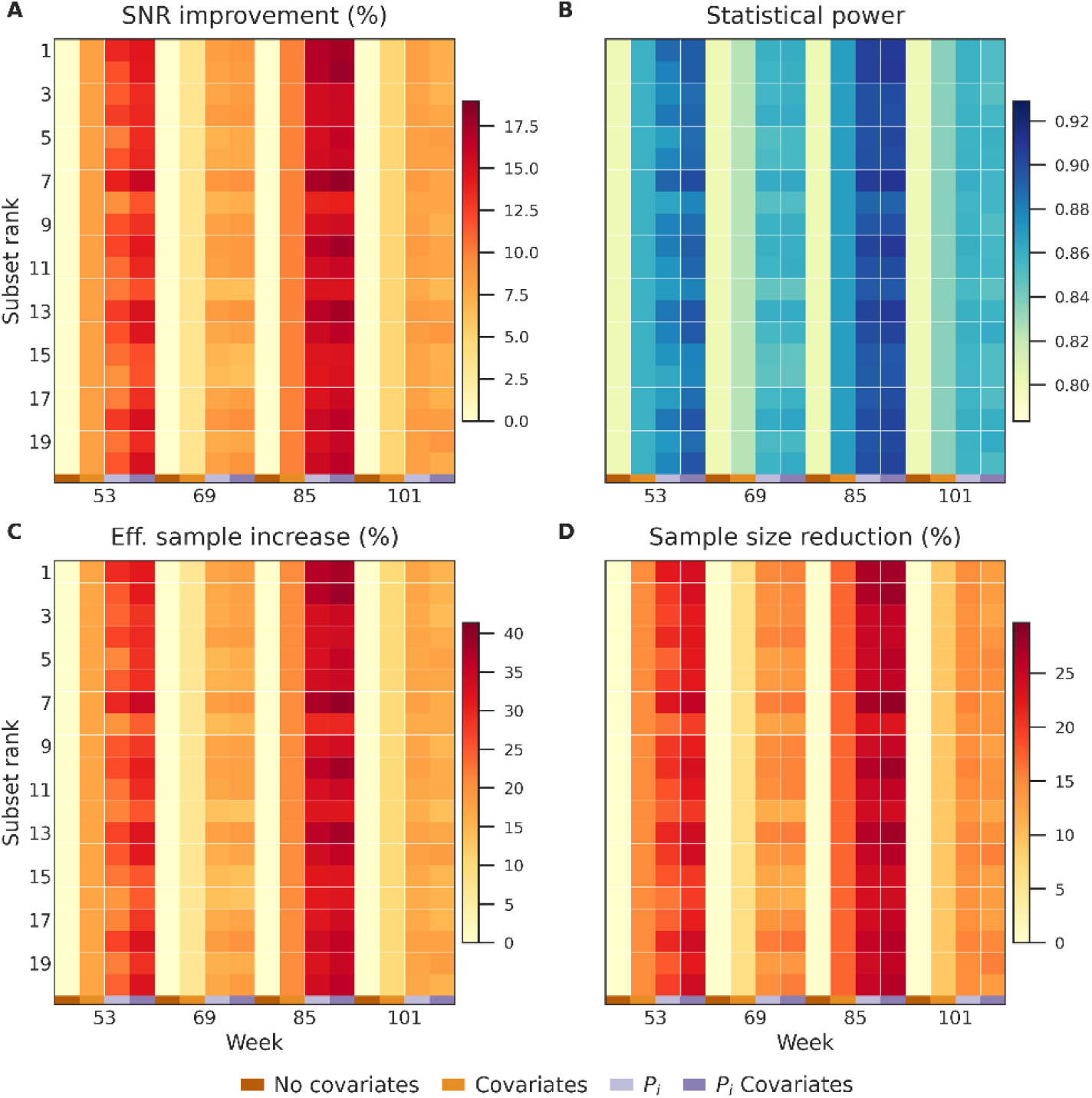
Impact of prognostic covariate adjustment on trial efficiency for TFC. Same as Figure 4, but for all 20 top performing 6-covariate TFC models.

## References

1. Andrews, D., Golchi, S., & Collins, D. L. (2026). A digital twin methodology using retrospective patient data for sample size reduction in Alzheimer’s disease clinical trials. Alzheimer’s Research & Therapy, 18(1), 2024.

2. Andrew, S. E., Paul Goldberg, Y., Kremer, B., Telenius, H., Theilmann, J., Adam, S., … & Hayden, M. R. (1993). The relationship between trinucleotide (CAG) repeat length and clinical features of Huntington’s disease. Nature genetics, 4(4), 398–403.

3. Angelopoulos, A. N., Bates, S., Fannjiang, C., Jordan, M. I., & Zrnic, T. (2023). Prediction-powered inference. Science, 382(6671), 669–674.

4. Boareto, M., et al. (2025). Modeling disease progression and placebo response in Huntington disease: insights from Enroll-HD and GENERATION HD1 cohorts. Neurology, 104(10), e213646.

5. Devanarayan, V., Ye, Y., Zhu, L., Tian, L., Kramer, L., Irizarry, M., & Dhadda, S. (2025). Predicted natural progression as an Alzheimer’s prognostic covariate improves the precision of lecanemab efficacy assessments and clinical trial efficiency. Alzheimer’s & Dementia, 21(3), e70045.

6. Ghazaleh, N., Houghton, R., Palermo, G., et al. (2021). Ranking the predictive power of clinical and biological features associated with disease progression in Huntington’s disease. Frontiers in Neurology.

7. Kieburtz, K., et al. (1996). Unified Huntington’s disease rating scale: reliability and consistency. Movement Disorders, 11(2), 136–142.

8. Ko, J., Furby, H., Ma, X., et al. (2022). Clustering and prediction of disease progression trajectories in Huntington’s disease: An analysis of Enroll-HD data using a machine learning approach. Frontiers in Neurology.

9. Landwehrmeyer, G. B., Fitzer-Attas, C. J., Giuliano, J. D., Gonçalves, N., Anderson, K. E., Cardoso, F., … & Squitieri, F. (2017). Data analytics from Enroll-HD, a global clinical research platform for Huntington’s disease. Movement Disorders Clinical Practice, 4(2), 212–224.

10. Langbehn, D. R., Hayden, M. R., Paulsen, J. S., & PREDICT-HD Investigators of the Huntington Study Group. (2010). CAG-repeat length and the age of onset in Huntington disease (HD): a review and validation study of statistical approaches. American Journal of Medical Genetics Part B: Neuropsychiatric Genetics, 153(2), 397–408.

11. McColgan, P., et al. (2023). Tominersen in adults with manifest Huntington’s disease. New England Journal of Medicine, 389(23), 2203–2205.

12. Mohan, A., Sun, Z., Ghosh, S., et al. (2022). A machine-learning derived Huntington’s disease progression model: insights for clinical trial design. Movement Disorders.

13. Poulet, P. E., Tran, M., Tezenas du Montcel, S., Dubois, B., Durrleman, S., & Jedynak, B. (2025). Prediction-powered inference for clinical trials: application to linear covariate adjustment. medRxiv.

14. Schuler, A., et al. (2022). Increasing the efficiency of randomized trial estimates via linear adjustment for a prognostic score. The International Journal of Biostatistics, 18(2), 329–356.

15. Sun, Z., Ghosh, S., Li, Y., et al. (2019). A probabilistic disease progression modeling approach and its application to integrated Huntington’s disease observational data. JAMIA Open.

16. Sun, W., Zhou, D., Warner, J.H., et al. (2020). Huntington’s disease progression: A population modeling approach to characterization using clinical rating scales. The Journal of Clinical Pharmacology.

17. Tabrizi, S.J., Schobel, S., Gantman, E.C., et al. (2022). A biological classification of Huntington’s disease: the Integrated Staging System. The Lancet Neurology.

18. Tabrizi, S. J., Scahill, R. I., Owen, G., Durr, A., Leavitt, B. R., Roos, R. A., … & Langbehn, D. R. (2012). Potential endpoints for clinical trials in premanifest and early Huntington’s disease in the TRACK-HD study: analysis of 24 month observational data. The Lancet Neurology, 11(1), 42–53.

19. Tran, M., Poulet, P. E., Petit, E., Durr, A., Klockgether, T., Ashizawa, T., Coarelli, G., & Tezenas du Montcel, S. (2026). How to improve statistical power in a trial with SCA2 patients using natural history data. Trials, 27(1), 297.

20. US Food and Drug Administration (FDA). Adjusting for covariates in randomized clinical trials for drugs and biological products: guidance for industry (May 2023), https://www.fda.gov/regulatory-information/search-fda-guidance-documents/adjusting-covariates-randomized-clinical-trials-drugs-and-biological-products.

21. Warner, J. H., et al. (2022). Standardizing the CAP Score in Huntington’s Disease by Predicting Age-at-Onset. Journal of Huntington’s Disease, 11(2), 153–171.

22. Wijeratne, Peter A., et al. (2021). Revealing the timeline of structural MRI changes in premanifest to manifest Huntington disease. Neurology: Genetics 7.5: e617.

